# Oral health and its association with self-rated health in Colombian community-dwelling older adults

**DOI:** 10.1101/2022.02.24.22271460

**Authors:** Luis Carlos Venegas-Sanabria, María Manuela Moreno-Echeverry, Miguel German Borda, Diego Andrés Chavarro-Carvajal, Carlos Alberto Cano-Gutierrez

## Abstract

**Objective:** Determine the association between different parameters of oral health and the self-rated health (SRH) in Colombian community-dwelling older adults.

**Methods:** This is a secondary analysis of the SABE-Colombia study performed in 2015. The dependent variable was defined as the SRH status assessed by the question “Compared with other people your age: Do you consider your health status to be: better, equal or worse?” The oral health parameters were edentulism, the GOHAI score and the use of dental prosthesis. Three multivariable logistic regression was performed compared better vs. worse, better vs. equal and equal vs. worse health status, adjusted by socio-demographic variables, comorbidities, and geriatric syndromes.

**Results:** After the exclusion of missing data, 18180 persons were included in the final analysis. 10.6, 37.6 and 51.6% persons reported worse, equal and better SRH, respectively. The worse SRH group had a higher proportion of > 80 years old persons, dependence, cognitive impairment, and depressive symptoms. The presence of edentulism and a low score of GOHAI were more frequent in the worse SHR group. After the multivariable logistic regression, all parameters of oral health status were associated with a worse SRH.

**Conclusion:** In our study, the oral health parameters were associated with self-rated health status. This result supports the inclusion of oral health in comprehensive geriatric assessment.

## Introduction

Due to the demographic transition and current advances in medicine, the older people’s population is steadily increasing^1^. Consequently, chronic pathologies have risen their prevalence, a situation that carries an increase in functional impairment, the need for caregivers. This situation forces to better health services and public policies. However, together with the rise of chronic pathologies the prevalence of other conditions that traditionally have been incorrectly considered as normal in older adults is also increasing ^2^. Among them, poor oral health and lost teeth have been shown to compromise the overall health and quality of life of adults and, most especially, older adults ^3,4^.

According to the World Health Organization, up to 30% of the world population between 65 and 74 years old, does not have natural teeth, percentages that are higher in low and middle-income countries ^5^. In general terms, we can define edentulism as the loss of teeth, that can be partial or total. The importance of edentulism lies in that affects the well-being of individuals, increases the risk of infections, alters the deglutition process, generates speech problems, social isolation, and malnutrition, just to mention a few of their consequences. ^6,7^ Research has shown that there is a direct relationship between the number of lost teeth, poor self-rated health and poor quality of life ^2,8,9^.

However, have natural teeth is not the only component of oral health^5^. Factors such dental caries, periodontal disease, xerostomia, specific pathologies as oral cancer, and the wrong use of dental prostheses ^8,10–12^, can deteriorate the oral health, and consequently, the overall health status and the quality of life^13^. Negative consequences of the oral health problems on older adult’s health condition are present regardless of functional status, cognitive impairment, poor social-economic conditions, or institutionalization. Further, preexisting health conditions can be exacerbated by any of these oral health problems.^10,14,15^

Due to the importance of oral health in the older adults, some instruments have been developed to assess the oral state and the impact of poor oral health. One of these instruments is the Geriatric Oral Health Assessment Index (GOHAI) ^16^, an index used to assess the oral health-related quality of life (OHRQoL). GOHAI measures three items: functionality (chewing, swallowing and speaking), the psychosocial aspect (discomfort when talking and physical appearance) and whether the pain is present when talking or eating. The Spanish version of the GOHAI has acceptable psychometric properties ^17^, it has been used in community-dwelling older adults and was validated in Colombian population. ^18,19^

The aim of this paper is to determinate the association between oral health-related quality of life, the present of natural teeth, and the use of dental prostheses (fixed or removable), with the self-rated health status in Colombian community-dwelling older adults.

## Materials and methods

### Setting and participants

#### Design of the survey

We analyzed data from the SABE (Salud, Bienestar y Envejecimiento-Health, Wellbeing, and Aging) 2015 Colombia study^20^, which is a cross-sectional study that included 23.694 community-dwelling subjects aged 60 years or more, from both rural and urban areas in Colombia. The sample design met the following characteristics: (1) regional representative; (2) self-representation of the country’s most important regions; (3) urban and rural sample size representation; (4) stage selection in accordance with municipal cartography. For the calculation of the sample size an estimated minimum proportion (P) of 0.03, a design effect (Deff) of 1.2, a relative standard error (Esrel) of 0.05 and a percentage of non-respond of 20% were considering. The survey had three components: (1) A questionnaire, which covered active aging determinants such as anthropometry, blood pressure measurement, physical function, and biochemical and hematological measures; (2) a subsample survey among family caregivers; (3) a qualitative study with gender and cultural perspectives of QoL in order to understand different dimensions of people. The instrument used in the SABE Colombia study were derived from the international instruments designed for the original SABE study, conducted in five Latin-American capital cities between 1999 and 2000. However, it was modified and adapted to Colombian context. All the instruments and scales were validated for Colombia ^20^. The first part of the questionnaire was the cognitive evaluation with the Mini-Mental State Examination Short Form (MMSE-SF). People with cognitive impairment according MMSE-SF (17.5%) required of a proxy to complete the survey. The other sections of the survey were: socioeconomical status, social network, housing and environment, social activities and hobbies, displacement and internal migration, nutritional status and behaviors, cognition and affect, daily life activities, health status and medical conditions, and anthropometry. The SABE Colombia study was executed from 2014 to 2015 by research groups of the Universidad del Valle and the Universidad de Caldas, with the operational support of the National Consulting Center (CNC) for fieldwork. The Colombian Ministry of Health and Colciencias (the Colombian Agency of Science) funded the study contract code 764-2015. Ethics committees of both University of Caldas and University of Valle reviewed and approved the SABE study protocol. All participants provided informed consent to participate in the survey.

#### Data and participants used in the secondary analysis

From the total SABE Colombia sample (n = 23,694), we excluded the participants that required a proxy (n = 4,690) to reduce confounding factors, as can be the impact of cognitive impairment in the self-rated health status. We also excluded the participants with incomplete data about oral health (n = 824). Finally, 18,180 subjects were included in the analysis. We used an anonymized version of the data base.

The present study was a secondary analysis of SABE Colombia study, and the ethics and scientific committee of both the Aging Institute at Pontificia Universidad Javeriana and Hospital Universitario San Ignacio approved, with the identification number 2017/180.

### Variables

#### Dependent variable

We used Self-Rated Health status as the dependent variable. It was defined by the question “*Compared with other people your age: Do you consider your health status to be: better, equal or worse?*” Considering the self-rated health status as a continuum, we decided to analyze the different states from each other. For this purpose, we compared the three possible answers in three different analysis: (1) better vs. worse; (2) better vs. equal; (3) equal vs. worse. We decided to use the self-rated comparison with other people instead of the comparison with oneself in the past time because we wanted to reduce the risk of memory bias.

#### Independent variables

As independent variables, we used three self-reported parameters to evaluate oral health: number of natural teeth, the Geriatric Oral Health Assessment Index (GOHAI) and the use of a prosthetic device. Other variables related to oral health, as dentist treatments, oral hygiene habits, or self-rathe oral health status, did not been included in the survey. Number of natural teeth was assessed by the question: “*Do you have natural teeth on the top/bottom dental arch?” Possible answers were: “Yes, from 1 to 5; Yes, from 6 to 10; Yes, 11 or more; No, if do not have any*.” We consider included in the analysis participants without natural teeth on the top and bottom dental arch (total edentulism), and participants with more of 22 teeth in total. The GOHAI^16^ was used to evaluate the oral health-related quality of life (OHRQoL). It includes the measures of three items: oral function (chewing, swallowing, and speaking), psychosocial aspects (discomfort when talking and physical appearance) and pain (when talking or eating). Scores of less than 57 indicate poor OHRQoL. The Spanish version has an acceptable psychometrics propriety and has been used in community-dwelling older adults. The use of prosthesis was assessed by the question: “*Do you have any of the following dental prostheses in the upper/lower part of your mouth (Fixed prosthesis, Partial removable prosthesis, Total removable prosthesis, or Implants)?*” To the analysis, we considered any positive answer about the use of prosthesis.^17–19^

#### Covariates

The age was categorized into three groups: 60-69, 70-79 and ≥80 years. Depressive symptoms were evaluated with the Geriatric Depression Scale created by Yesavage^21^ and were defined as a total score of 10 or more. The basic activities of daily living (feeding, moving from chair to bed and return, doing personal toilet, getting on and off toilet, bathing self, walking on a level surface, ascending and descending stairs, dressing and undressing, continence of bowel, controlling bladder) was assessed through the Barthel index^22^. We considered as functional independence a score of 90 or high^23^. Due to an exclusion criterion was the presence of significant cognitive impairment, we used as confounding variable the mild cognitive impairment defined as a score of Clinical Dementia Rating (CDR)^24^ of 0.5. Frailty, a pre-disability state,^25^ was assessed using the FRAIL scale,^26^ which assesses five items: fatigue, resistance, aerobic capacity, number of illness, and loss of weight. Each item equals one point, and frailty was considered with a score ≥ 3. The presence of chronic diseases was evaluated by asking participants if they had been previously diagnosed with mental illness, diabetes mellitus (DM), Chronic obstructive pulmonary disease (COPD), cardiovascular disease (hypertension, cerebrovascular disease or coronary disease), osteoarticular diseases (arthrosis, rheumatoid arthritis, osteoporosis or rheumatism) or cancer (any type). Considering the effect that could have over the self-rated health status, general symptoms were evaluated through self-reported (dyspnea, dizziness, back pain, weakness, exhaustion, nausea and vomiting, joint pain, and insomnia). We recorded these variables through dichotomic answers (yes or no).

#### Statistical analysis

The SABE Colombia sample design was explained previously (see setting and participants). Initially, we used univariate analyses to explore extreme values and assess for normal distribution, as well as to adjust and categorize variables. Regarding descriptive statistics, categorical variables were presented through frequencies (absolute and relative). For continuous variables, we used the Kolmogorov-Smirnov statistic to determine the normal distribution of the variables. The variables with normal distribution were reported through means and standard deviations (SD), and for variables that were non-normally distributed the median and inter-quartile ranges (IQR) were reported. Then, an analysis was performed in order to contrast the difference between the three SRH groups (better, equal, and worse). Chi-square tests were used for categorical variables and t-tests were employed for continuous variables. For the regression analysis, we considered three different models. In the first model, we compared the participants that reported a better SRH with the participants that reported it worse. The next model compared participants with better vs. equal SRH and the third model compared the ones with equal vs. worse SRH. A multivariate logistic regression considering the independent variables (the presence or absence of natural teeth, GOHAI score less than 57, and the use of a prosthesis) was performed. To each model, we performed an adjusted analysis looking to find the most parsimonious model including socioeconomic, functional, affective, of medical conditions, and of the presence of general symptoms variables. The results were present as Odd Ratio (OR), and 95% confidence interval (95% CI). The statistical level of significance was set at p <0.05. Data were analyzed employing the statistic software SPSS® 24 (IBM® Statistics).

## Results

The descriptive analysis and the differences between the three SRH status groups are shown in table 1. Compared with other same-age persons, 10.6% reported worse SRH. 51.6% and 37.6% reported better or equal SRH respectively. In all groups, the female-proportion was high that the male (better = 56.8%, equal = 53.8%, worse = 59.1%). The median age in the SRH better group was 68 years (IQR 63-74), in the SRH equal group 68 years (IQR 63-74), and SRH worse group 68 years (IQR 64-75) (p = 0.01). In terms of distribution by age, we only found a significant difference in the subjects older than 80 years (better = 9.8%; equal = 9.6%; worse = 12.3). Regarding to living in a rural area, we found a difference between the three groups (better = 21.2%, equal = 29.1%, worse = 35.4%; p = 0.00). Non-difference in the proportion of persons cataloged as low economic income was found. The group of worse SRH had less functionality to BADL (88% vs. 97.8% and 96.9%). In addition, this group had a worse cognitive status and more depressive symptoms (30% and 9.6% respectively).

**Table 1.**
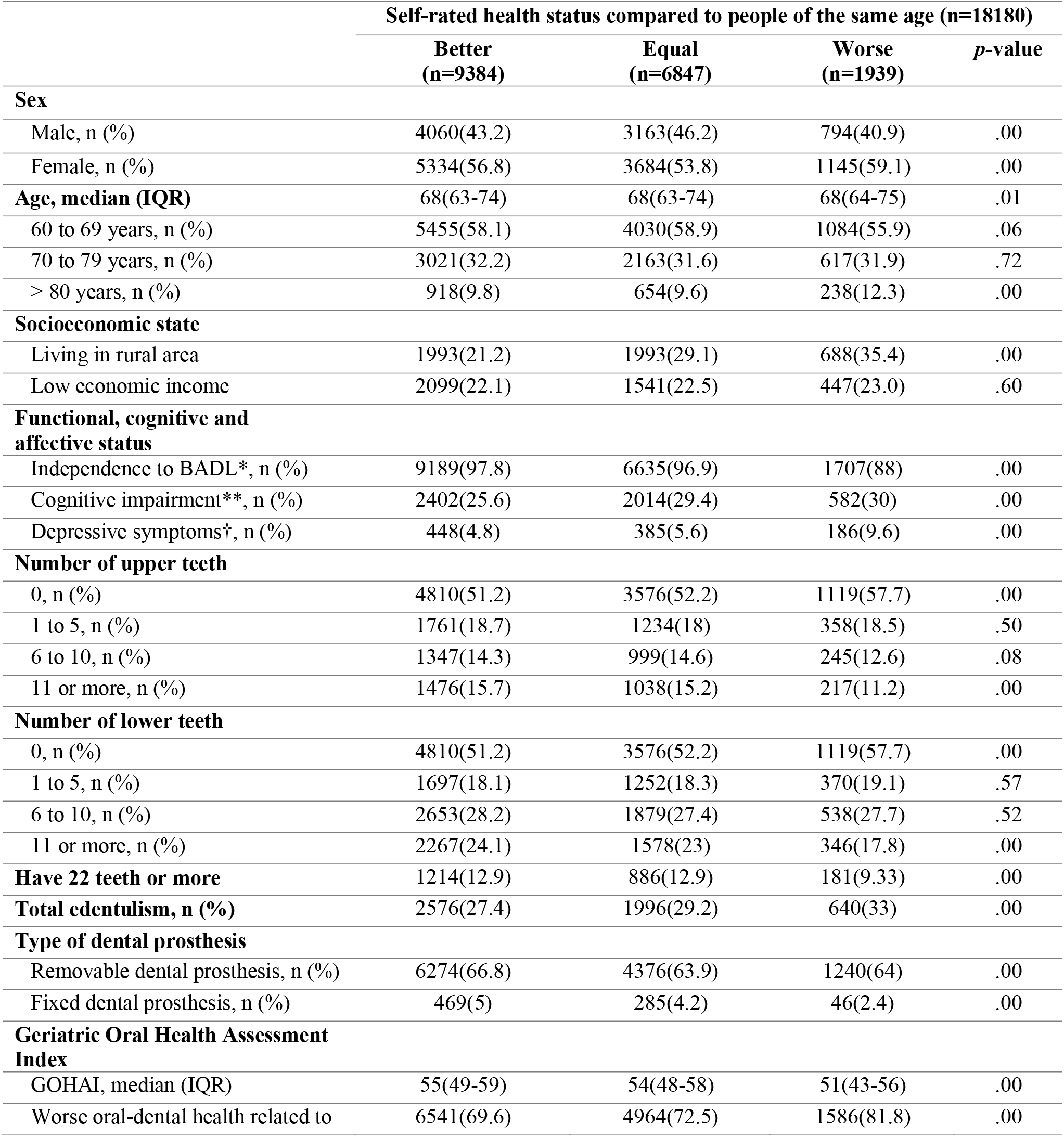
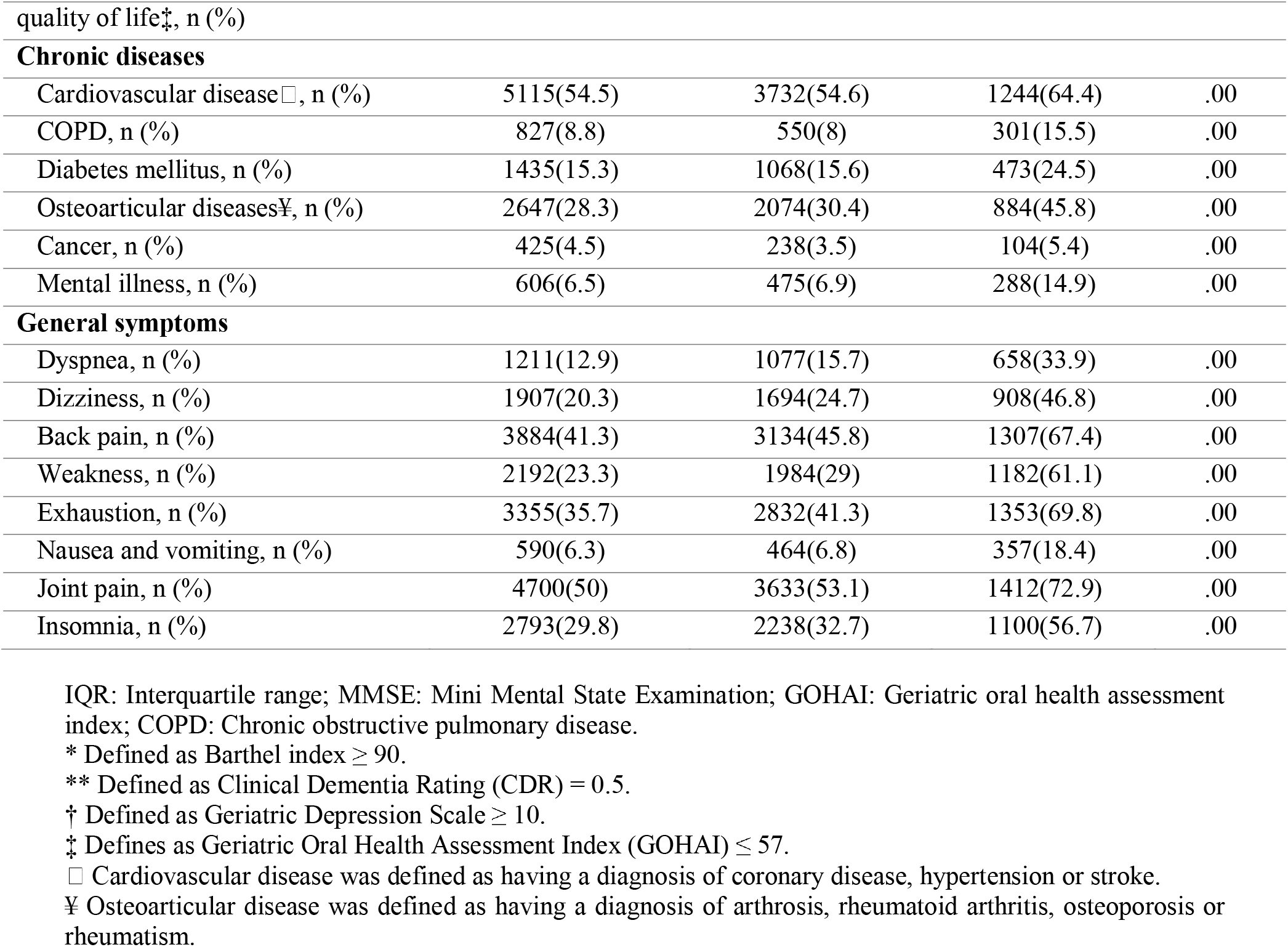
Bivariate analysis: Self-rated health status and Oral health.

The subjects with worse SRH had a higher prevalence of edentulism compared to those who reported a better or equal SRH (33, 27.4 and 29.2% respectively). The use of fixed and removable dental prosthesis was more frequent in the better SRH group (fixed = 5%; removable = 66.8% respectively) compared with the other two groups (equal = 4.2 and 63.9%; worse = 2.4 and 64%). The median of the GOHAI was 55 in the better SRH group, 54 in the equal and 51 in the worse (p = 0.00). The prevalence of worse oral-dental health related to the quality of life, defined by a GOHAI score of less than 57, was high in the three groups (better = 69.6%; equal = 72.5%, worse = 81.8%; p = 0.00). The prevalence of all chronic diseases and general symptoms were higher in the worse SRH group.

The multivariate logistic regression (table 2) was developed considering three different analysis (better vs. worse; better vs. equal; equal vs. worse) contrasting the three groups of SRH, taking as independent variables the presence of total edentulism, have 22 or more teeth, the use of a fixed or removable dental prosthesis, and the GOHAI less than 57. The three multivariate logistic regressions were adjusted using the forward stepwise selection variables method. The fist model (better vs. worse) was adjusted by age ≥ 70y, female, living in a rural area, diabetes, mental disease, osteoarticular diseases, dyspnea, dizziness, back pain, weakness, exhaustion, nausea and vomiting, joint pain, insomnia, frailty, functional independence, and depressive symptoms. The second one (better vs. equal) by age ≥ 70y, living in a rural area, female, osteoarticular disease, dizziness, exhaustion, weakness, functional independence, and depressive symptoms. Finally, the last model (equal vs. worse) was adjusted by age ≥ 70y, female, living in a rural area, diabetes, mental disease, osteoarticular diseases, dyspnea, dizziness, weakness, exhaustion, nausea and vomiting, joint pain, insomnia, frailty, functional independence, and depressive symptoms. In the first analysis, we found association between among all independent variables. GOHAI and total edentulism were associated with a worse SRH (OR 1.19; 95% CI 1.04-1.37 and OR 1.19; 95% CI 1.04-1.35, respectively), while the use of a fixed or removable prosthesis and presence of 22 natural teeth or more was associated with a better SRH (OR 0.59; 95% CI 0.42-0.82; OR 0.75; 95% CI 0.65-0.86; OR 0.80; 95% CI 0.65-0.98, respectively). In the second analysis, better vs. equal SRH, we found that the GOHAI less than 57 and the presence of 22 natural teeth or more did not have an association with the SRH. The use of a fixed (OR 0.84; 95% CI 0.72-0.98) and the use of a removable dental prosthesis (OR 0.86; 95% CI 0.80-0.93) was associated with have a better SRH, and the presence of total edentulism was associated with having an equal SRH compared with same age people (OR 1.13; 95% CI 1.05-1.22). In the third analysis, we found an association between the use of fixed or removable prosthesis and presence of 22 natural teeth (OR 0.61; 95% CI 0.59-0.88; OR 0.82; 95% CI 0.72-0.94; OR 0.72; 95% CI 0.59-0.88, respectively) and have an equal SRH.

**Table 2.**
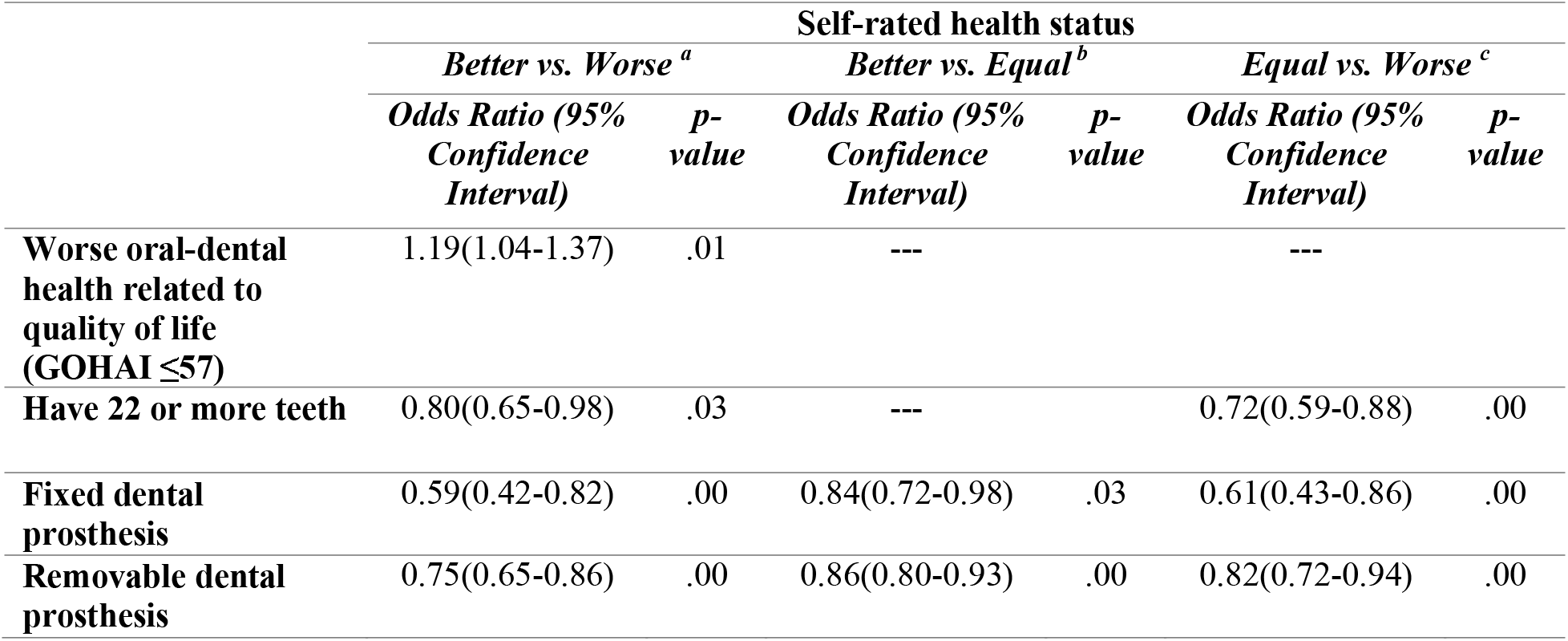
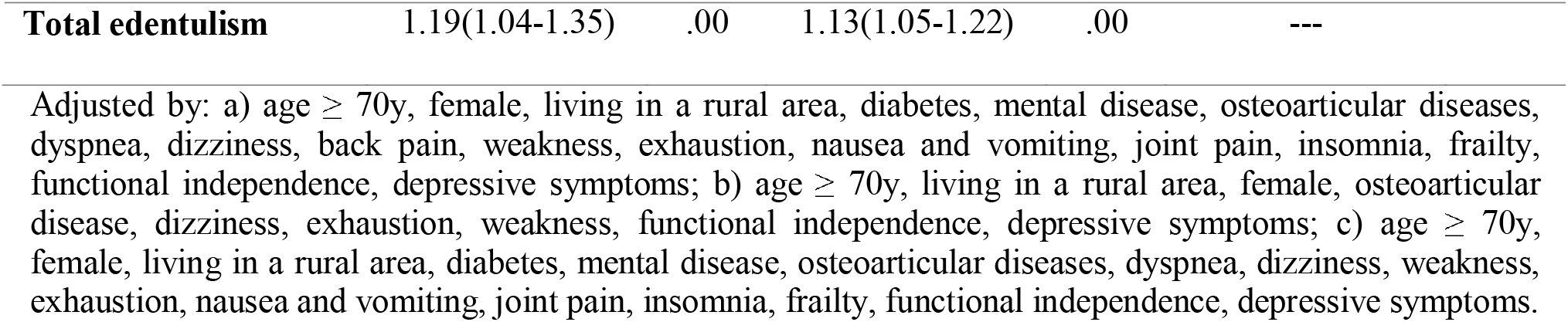
Adjusted multivariate logistic regression for self-rated health status and oral health.

|  | Self-rated health status |  |  |  |  |  |
| --- | --- | --- | --- | --- | --- | --- |
|  | Better vs. Worse <sup>a</sup> |  | Better vs. Equal <sup>b</sup> |  | Equal vs. Worse <sup>c</sup> |  |
|  | Odds Ratio (95% Confidence Interval) | p-value | Odds Ratio (95% Confidence Interval) | p-value | Odds Ratio (95% Confidence Interval) | p-value |
| **Worse oral-dental health related to quality of life (GOHAI $\leq 57$)** | 1.19(1.04-1.37) | .01 | --- |  | --- |  |
| **Have 22 or more teeth** | 0.80(0.65-0.98) | .03 | --- |  | 0.72(0.59-0.88) | .00 |
| **Fixed dental prosthesis** | 0.59(0.42-0.82) | .00 | 0.84(0.72-0.98) | .03 | 0.61(0.43-0.86) | .00 |
| **Removable dental prosthesis** | 0.75(0.65-0.86) | .00 | 0.86(0.80-0.93) | .00 | 0.82(0.72-0.94) | .00 |

|  |  |  |  |  |  |
| --- | --- | --- | --- | --- | --- |
| **Total edentulism** | 1.19(1.04-1.35) | .00 | 1.13(1.05-1.22) | .00 | --- |

Adjusted by: a) age ≥ 70y, female, living in a rural area, diabetes, mental disease, osteoarticular diseases, dyspnea, dizziness, back pain, weakness, exhaustion, nausea and vomiting, joint pain, insomnia, frailty, functional independence, depressive symptoms; b) age ≥ 70y, living in a rural area, female, osteoarticular disease, dizziness, exhaustion, weakness, functional independence, depressive symptoms; c) age ≥ 70y, female, living in a rural area, diabetes, mental disease, osteoarticular diseases, dyspnea, dizziness, weakness, exhaustion, nausea and vomiting, joint pain, insomnia, frailty, functional independence, depressive symptoms.

## Discussion

The results of this study suggest that a poor oral health (total edentulism and GOHAI<57), is associated with a worse SRH in the elderly. The presence of total edentulism was statistically associated with a worst SRH, similarly to what have been described by other authors. For example Tyrovolas et al., who found an association of edentulism with poor SRH (OR 1.38, 95% CI□=□1.03–1.83) and also depression^27^. In other Latin American countries this association had already been described; Borda et al found a significant association between edentulism and SRH with an incremental risk according to the number of missing teeth^2^. This means that having fewer teeth puts older people at greater risk of having poor SRH, and hence poor health in general. On the other hand, we found that had 22 natural teeth or more was associated with better SRH status in the three models. This confirms the importance that has to older people have the preservation of their natural teeth. In our study, a GOHAI<57 was associated with a worse SRH only when was compared with a better SRH compared with same age people, a relationship that has been described by other authors in different populations^6,28^.

This association maybe mainly a consequence of the impaired dentition on dietary restriction, food taste alteration, the need of food selection (consistence i.e.), different food preparations and food eating patterns. In fact, Locker et al^29^ found that 39% of edentulous elders were prevented from eating foods they would like to eat, 20% reported a decline in their enjoyment of food, and 14% avoided eating with others. Joshipura el al^30^ reported that when compared to dentate individuals, edentulous individuals consumed fewer vegetables, less fiber, and less carotene, and consumed more cholesterol and saturated fats.

Using historical records of oral health may be helpful to draw better health care plan for geriatrics population. Remarkably, we found that fixed dental protheses (FDP) were associated with a better SRH in the three groups analyzed, and nevertheless the removable dental protheses (RDP) showed a benefit in two of the three groups, the benefit was greater with the use of fixed protheses. A similar finding was described by Klotz et al ^31^, reporting that participants with FDP had the highest Oral Health Related Quality of Life (OHRQoL) when compared to patients with RDP. Even though some studies have described a less use of FDP due to post operatory pain and cost, other studies investigating masticatory performance revealed that chewing efficiency decreases from patients wearing FDP to those wearing RPD^32^, and in general clinicians (using their perception of objective criteria) view fixed prostheses as being superior to removable prostheses, both functionally and aesthetically. An important aspect of considering is the cost of both removable and fixed prostheses. In general, the FDP has a higher cost, with which low-income people may not have access to this kind of prostheses.

It is important to highlight that in our knowledge, this is the first study that takes oral health broadly, including within the variables of our analysis, the number of teeth, total edentulism, the use of removable or fixed dental prostheses and the GOHAI score at the same construct. This perspective enriches the analysis of the data obtained and its relationship with the SRH, which allows us to characterize better the oral health determinants that are associated with the SRH of the people in our country.

There are some limitations to our study; first, the cross-sectional design limits the potential for etiological conclusions. Second, residual confounding may exist since we could not adjust for factors such as dietary habits, social cohesion, access to dental care due to lack of data. Thus, their confounding and individual effects remain unknown. One the other hand our study has several strengths, to the best of our knowledge, this is the first study in our country to evaluate the impact of oral health in the self-rated health status, and our are important for evaluating interventions to improve the health status in older adults and strategies to limit the health consequences of poor oral conditions.

In conclusion, in our study, the parameters of oral health, the oral health-related quality of life measured by GOHAI, the presence of the total edentulism, the number of natural teeth, and the use of the dental prosthesis, were associated with the self-rated health status in older Colombian persons. The association was higher when the persons with better were compared with persons with worse self-rated health status. Prospective studies are required to confirm this association.

## Data Availability

All data produced in the present work are contained in the manuscript

https://github.com/lcvenegas/srh_or

